# Evaluating the economic and health impact of proactive genomic epidemiology in a hospital setting

**DOI:** 10.1101/2025.01.30.25321399

**Authors:** Frederik Boetius Hertz, Karen Leth Nielsen, Dmytro Strunin, Jelena Erdmann, Martin Lucas Jørgensen, Theiss Bendixen, Roshkan Srinathan, Rasmus L. Marvig, Steen Christian Rasmussen, Asger Nellemann Rasmussen, Christian Salgaard Jensen, Jenny Dahl Knudsen, Susanne Häussler

## Abstract

**Background:** Genomic epidemiology which combines whole genome sequencing (WGS) of bacterial isolates with patient movement data, promises improved detection, prevention, and management of infection transmission, enhancing patient safety and reducing healthcare costs. However, evidence on its cost-effectiveness and clinical utility remains limited, as initial studies were restricted to selected pathogens and based on extrapolated assumptions from partial WGS data.

**Methods:** We conducted a 28-month observational study at Rigshospitalet, a 1 200-bed tertiary hospital in Copenhagen. The study involved collecting patient movement and WGS data for clinical isolates of 19 bacterial species, regardless of their site of isolation, or antibiotic susceptibility profile. This included sequencing of 18 940 clinical isolates from 7 760 patients.

**Findings:** We found that 27·1 % of culture-positive hospitalized patients harboured a pathogen genetically related to another patient’s isolate. 69 % had an epidemiological link, indicating transmission between patients, with *Enterococcus faecium* being the most prevalent. Notably, there were 2·2 times more transmissions of antibiotic-susceptible than resistant isolates. We estimated that prevention based on genomic epidemiology could potentially generate net savings of €1·25 million annually and avoid more than 750 disability adjusted life years (DALYs).

**Interpretation:** Our holistic, proactive genomic epidemiology approach reveals previously unexplored transmission landscapes. We discovered that transmission is widespread, varies significantly between species, and is not limited to resistant isolates. Our results highlight the potential for WGS-informed infection control, greater savings by including susceptible isolates, and an incremental cost-effectiveness ratio (ICER) classification by pathogen species.

**Funding:** Novo Nordisk Foundation, Beta.Health, Den Frie Forskningsfond

## Introduction

Healthcare-associated infections (HAIs) are among the most common complications of hospital care (1–3). If not adequately treated, HAIs cause suffering, incapacity and death, and impose an enormous financial burden on healthcare systems as well as on society in general (2,4–6). The European Centre for Disease Prevention and Control (ECDC) estimates that HAIs in Europe affect over 3·5 million people annually, causing more than 90 000 deaths and about 2·5 million disability-adjusted life years (DALYs) (7). In Denmark, HAIs affect 7-10% of admissions, with surgical wound infections alone accounting for an estimated 2% of the total Danish hospital costs in 2000 (8,9). Costs increase due to prolonged and extended antibiotic therapy, the need for advanced diagnostic tests or imaging techniques, more intensive medical treatments, including surgical procedures, and extended hospital stays (10–13). Furthermore, there are additional likely underestimated indirect costs, such as those incurred outside the hospital and costs borne by the patient (8). These statistics highlight the urgent need for the implementation of more effective measures to reduce HAIs. Morbidity, mortality and costs associated with HAIs caused by multidrug-resistant organisms (MDRO) are particularly high (4,5). Additionally, the increased use of broad-spectrum antibiotics, often required in these cases, further drives antibiotic resistance (14). As a result, the infection prevention and control (IPC) teams are closely monitoring MDROs in hospitals.

Genomic epidemiology, which combines the identification of genetically related bacterial isolates and patient movement data, is increasingly being used to detect transmission of MDROs (15,16). Knowledge on clonal variants of clinical isolates enables the refutation or confirmation of transmission and support the initiation of targeted IPC responses. However, not only antibiotic resistant, but also susceptible isolates are transmitted within hospitals. Along with bacteria from endogenous infection sources, transmitted susceptible isolates contribute, though largely unexplored, to high HAI rates (17).

Whole genome sequencing (WGS) technologies are increasingly being adopted as an effective alternative to conventional pathogen typing methods and have facilitated pathogen surveillance as well as rapid investigation and management of outbreaks (15,18–20). However, the successful integration of WGS in routine infection prevention and control relies on cost-effective workflows to generate affordable sequencing data. Furthermore, advanced analytics software is essential for timely interpretation of high-throughput genomic data (15,18,19). The effectiveness of a WGS-based surveillance tool depends heavily on the prompt identification of clusters of patients, who harbour genetically related isolates and the swift delivery of this information to the infection control team. Several studies have evaluated the benefits of large-scale bacterial infection surveillance, their role in detecting transmissions, and the impact of transmitted isolates on HAI rates. Such evaluations are crucial to support the broader application of prospective routine WGS and to estimate the health burden associated with the transmission of pathogens (15–17).

Here, we performed a prospective observational study and collected a total of 18 940 clinical isolates from 7760 unique patients in a tertiary hospital setting over a 28-month period. All isolates were subjected to WGS in a non-discriminatory approach and included a broad spectrum of gram positive and gram negative species, from various body sites, from both intensive care unit (ICU) and non-ICU patients, and regardless of their antibiotic resistance profile. Our goal was to estimate the potential for reducing transmission and the associated clinical and economic benefits. We discovered that 27·1 % of culture-positive hospitalized patients harboured a bacterial isolate closely genetically related to an isolate from another patient and found significant potential for cost savings through the implementation of genomic epidemiology for MDROs. Even greater overall savings could be possible by extending genomic epidemiology to include susceptible isolates, as the majority of previously undetected transmissions involve susceptible isolates from various species. Of note, the incremental cost-effectiveness ratio (ICER) varied significantly between bacterial species, with WGS applied to *Enterococcus faecium* providing the highest return on investment.

## Methods

### Setting

Copenhagen University Hospital - Rigshospitalet is a tertiary hospital with 1 200 beds, situated in Copenhagen, Denmark, predominantly used for referral and highly specialized treatments.

### Data sources

In Denmark, all citizens are assigned a unique civil registration number (CPR number) at birth or upon immigration, enabling individual-level linkage of nationwide registers. The laboratory information system MADS (Mads-group, University Hospital Aarhus, Denmark) (21) was used to extract information about all clinical samples send for culturing at the Department of Clinical Microbiology (DCM) at Rigshospitalet. The data included the following variables: microbiological sample identifier (unique number), dates of receiving the sample, collection date, date of delivering the test results, test category (e.g., focus of infection, screening for MDRO, clinical sample), species of identified bacteria and their categorisation into MDRO or non-MDRO based on phenotypic resistance testing. Furthermore, clinical metadata were extracted from the patient file system, the EPIC^TM^ platform called Sundhedsplatformen (SP) (21). These included department codes for admission or medical encounter, patient admission type (inpatient, outpatient, and others), date of admission, date of discharge, and patient identifier codes, which were used to match data points (instances) across the different datasets.

### WGS-based surveillance of hospital transmission

MDROs were defined based on the Dutch nosocomial infection surveillance guidelines (22,23). The MDROs of this study included Methicillin-resistant *Staphylococcus aureus* (MRSA), Vancomycin-resistant enterococci (VRE), Extended-spectrum beta-lactamase (ESBL)-producing Enterobacterales resistant to two additional classes of antibiotics (cephalosporins, fluoroquinolones and aminoglycosides), *Pseudomonas aeruginosa* isolates resistant to three classes of antibiotics, and Carbapenem-resistant gram negatives. As part of the infection surveillance system at Rigshospitalet, all MRSA, *Enterococcus faecium* VREs and carbapenemase producing organisms (CPOs) are collected and stored at the DCM, and subjected to WGS, with a routine turnaround time of 14 days. In this prospective observational study, additionally all clinical isolates belonging to the following species were sequenced: *Escherichia coli, Raoultella/Klebsiella* spp*., P. aeruginosa, Acinetobacter* spp*., Enterobacter* spp*., Citrobacter* spp*., Enterococcus* spp*., Serratia* spp., and *Stenotrophomonas maltophilia*, regardless of antibiotic susceptibility profiles, or focus of infection. For each species, the study included one isolate per patient, per sample type, per day.

### Sequencing and detection of clusters

Bacterial isolates were transferred into a 96-deep-well plate containing lysis buffer (1x TE buffer pH=7·8-8·2, 1% Triton-X) from an overnight agar plate culture and incubated first with lysostaphin (21µg/ml) (gram positive isolates only) and lysozyme (25mg/ml) (gram positive and gram negative isolates) for 30 min at 37°C, followed by incubation with proteinase K (0·4mg/mL) and AL buffer (Qiagen) for 30 min at 56°C, both steps with 550 rpm shaking. After incubation 200µL ethanol (99%) was added to each sample and transferred to a spin column plate (EZ-96 DNA filter plate with 96-well collection plates). The plate was centrifuged for 10 min at 6000 rpm. Washing was performed with 500µL of wash buffer 1 (25% 3M acetate buffer, pH=8·0, 75% ethanol) and centrifuged for 10 min at 6000 rpm followed by another wash step (10% 3M acetate buffer, pH=8·0, 90% ethanol) and centrifugation (15 min at 6000 rpm to dry the column). The DNA was eluted in nuclease free water by centrifugation (10 min at 6000 rpm). Libraries were produced following the Hackflex workflow (24) and libraries were sequenced on either a NextSeq 500 (96 libraries per sequencing run) or NovaSeq 6000 instrument (384 libraries per sequencing run) generating 150 base paired-end sequencing reads.

The WGS data were analysed using an in-house analysis pipeline that included trimming with bbmap/36.49, assembly with shovill/1.0.4., applying Spades/3.14.0 and MLST-typing by mlst (Seemann T, https://github.com/tseemann/mlst). Assemblies were excluded from the study if the genome consisted of more than 400 contigs or if the genome size did not correspond to that of the respective species. Finally, the pipeline identified close genetic relatives in a two-step analysis. In the first step, close relatives were identified based on gene sequence-based Jaccard similarity, followed by detailed SNP analysis on the related isolates using SKA2 (25). More specifically, the Jaccard similarity between all pairs of isolates within a species was calculated: J(A,B) = |(A∩B)/(A∪B)|. This value was obtained by dividing the number of genes with identical DNA sequence by the total of unique gene variants in both genomes. SNP similarities were then computed for isolate pairs with a Jaccard similarity ≥ 0·8 indicating that at least 80% of the combined gene pool shares identical gene sequences, using SKA2.

We identified clusters of two or more patients with genetically related isolates, using a cut-off of 20 SNPs between the isolates. There’s no single SNP cut-off universally applicable to all species, and species with high genetic diversity may require a higher SNP cut-off to detect meaningful patterns of relatedness, while species with low diversity may require fewer SNPs. By setting the cut-off at 20 SNPs, we accept that the proportion of false-positive genetic links may be higher in some species (e.g., *Enterococcus* spp. (26,27)) compared to others. To counteract this bias, we additionally searched for epidemiological links, meaning we analysed whether the patients had direct or indirect contact in the hospital. We assume that isolate pairs with a false-positive genetic link are less likely to have an identified epidemiological link.

Within a cluster all patients harboured at least one isolate exhibiting a genetic distance of 20 SNPs or less to at least one other isolate from another patient. To determine when and where transmission may have occurred within each of the patient clusters, we searched for temporal and spatial links between patients. A direct epidemiological link was defined as patient admission to the same department during an overlapping period of at least one day. An indirect epidemiological link was defined as admission to the same department within a maximum gap of 14 days.

### Analytic model

We aimed to estimate the cost-effectiveness of proactive WGS of HAI-associated bacterial pathogens combined with standard infection prevention and control practice, versus standard practice alone. Decision tree modelling was applied to analyse the economic implication of implementing genomic epidemiology at Rigshospitalet. This method maps out different choices and their potential outcomes, particularly in terms of costs and health outcomes based on an annual cohort of newly admitted, hospitalized patients and has been used to compare various treatments or interventions. The decision analytical model-design followed the approach previously described by Dymond et al. in 2019 in their cost-effectiveness study of genomic surveillance of MRSA (28). In the simulation model, patients can follow different “branches” of treatment and outcomes, such as MDRO-positive/negative, MDRO-infection/colonization and death/discharge. The different costs that were assigned to the patients in the different branches are listed in Table 1. Cost components that are associated with additional cost for an infection with an MDRO or a susceptible isolate as well as additional costs associated with the colonization with an MDRO were derived from the Danish Health Data authorities (https://sundhedsdatastyrelsen.dk/da/english/health_finance or https://sundhedsdatastyrelsen.dk/da/afregning-og-finansiering/gruppering-drg/interaktiv-drg), from the literature and DCM data. The cost parameters were restricted to a direct hospital perspective (e.g., extra cost per admission day) and were delivered without overhead (management and maintenance of buildings and hardware). We used the average admission time at Rigshospitalet of 4·6 days (in 2023) as baseline admission time (https://www.rigshospitalet.dk/english/about-us/Pages/facts-and-figures.aspx). Costs associated with MDRO colonization include costs to health professional personal protective equipment, microbiology tests, and cleaning. Patients who were infected with an MDRO (estimated via the finding of a pathogen in a clinical sample and not a screening sample) accrued these same costs, however, additional cost for prolonged length of stay in the hospital (LOS) were added. The mean prolonged LOS for each infected patient (MRDO or non-MDRO) was assumed to be 7·8 days based on published data (13). The cost per WGS includes library preparation and sequencing, laboratory technician time, and indirect overhead costs of 25%.

**Table 1.** Inputs for costs. ^⁕^ Taken into account the incidences of healthcare-associated (HA) pneumonia, HA primary bloodstream infection, HA urinary tract infection, surgical site infection, and HA neonatal sepsis, as well as the DALYs per case from the recent publication on the burden of HAIs in Europe (6), we calculated a cumulative burden of common HAIs to be 1·65 DALYs per case (95 % CI 1·45 – 1·85). Since, according to the study (6), an average of 75% of the DALYs can be attributed to YLL and 25% to YLDs, we estimated a cumulative burden of common HAIs to be 1·24 YLLs per case (95% CI 1·09-1·39) and 0·41 YLDs per case (95% CI 0·36-0·46).

| Parameter | Value | Source | Lower bound | Upper bound |
| --- | --- | --- | --- | --- |
| Excess length of hospital stay (LOS) attributable to HAI | 7·8 days | (13) | 5·7 (95% CI) | 9·9 (95% CI) |
| Costs for patients colonized with MDRO on a medical ward | 325 € x 4·6 days (1 495 €) | Danish Health Data authorities | 1 196 € (- 20%) | 1 794 € (+20 %) |
| Costs for patients infected with MDRO on a medical ward | 5 070 € | Danish Health Data authorities | 4056 € (-20%) | 6 084 € (+ 20%) |
| Cost of infection with non-MDRO | 2 535 € | Danish Health Data authorities | 2028 € (- 20%) | 3 042 € (+ 20%) |
| Relative fraction of MDRO versus non-MDRO among clinical isolates included in this study | 9·75 % | Based on data from this study |  |  |
| Cost per WGS (includes library preparation, sequencing laboratory technician time, and indirect overhead costs) | 35 Euro | DCM | 28 € (-20 %) | 42 € (+ 20%) |
| Cumulative burden of common HAIs, | 1·65 DALYs * | (6) | 1·45 (95% CI) | 1·85 (95% CI) |
| YLLs (years of life lost) | 1·24 YLLs * | (6) | 1·09 (95% CI) | 1·39 (95% CI) |
| YLDs (years lived with disabilities) | 0·41 YLDs * | (6) | 0·36 (95% CI) | 0·46 (95% CI) |

To estimate the clinical utility of a proactive WGS approach, we assumed that none of colonized patients would suffer from pathogen-related causes. However, we assigned a clinical burden of HAIs to infected patients based on published data (6). The clinical burden of HAIs has previously been estimated in the *Burden of Communicable Diseases in Europe* (BCoDE) project (6). The study accounted for the incidences of different HAIs and estimated years of life lost due to mortality (YLL), years lived with disability (YLD), and disability-adjusted life years (DALYs) for the most common HAIs. DALY represents a composite health measure that includes YLDs after onset of a disease and YLLs compared to a standardized life expectancy.

Sensitivity analyses were conducted to access the robustness of the overall results of the model and to quantify the effects of uncertainty on cost-effectiveness following the implementation of proactive WGS.

## Results

### Implementation of non-discriminatory WGS on the most frequent nosocomial pathogens processed in the Department of Clinical Microbiology

Rigshospitalet treats more than 300 000 unique patients every year, with an average of 10 670 colonized/infected patients (per year over 2022-2023). In this observational study, we included all patients, who were colonized/infected with a MRSA, as well as with isolates irrespective of their resistance profile of the following bacterial species: *E. coli, Raoultella/Klebsiella* spp*., P. aeruginosa, Acinetobacter* spp*., Enterobacter* spp*., Citrobacter* spp*., Enterococcus* spp*., Serratia* spp., and *S. maltophilia*. Overall, 18 940 bacterial isolates (one isolate per patient per day per isolation site) from 7 760 unique patients were included in the study. This corresponded to the sequencing of around 33·5 % of bacterial isolates processed at the DCM during the study period, considering one isolate per day per patient for each sample type. 27·4 % of the MDRO isolates included in this study were obtained from screens. The relative distribution of clinical isolates by bacterial species and of the patients colonized/infected with at least one unique isolate of these species is depicted in Figure 1.

**Figure 1:**
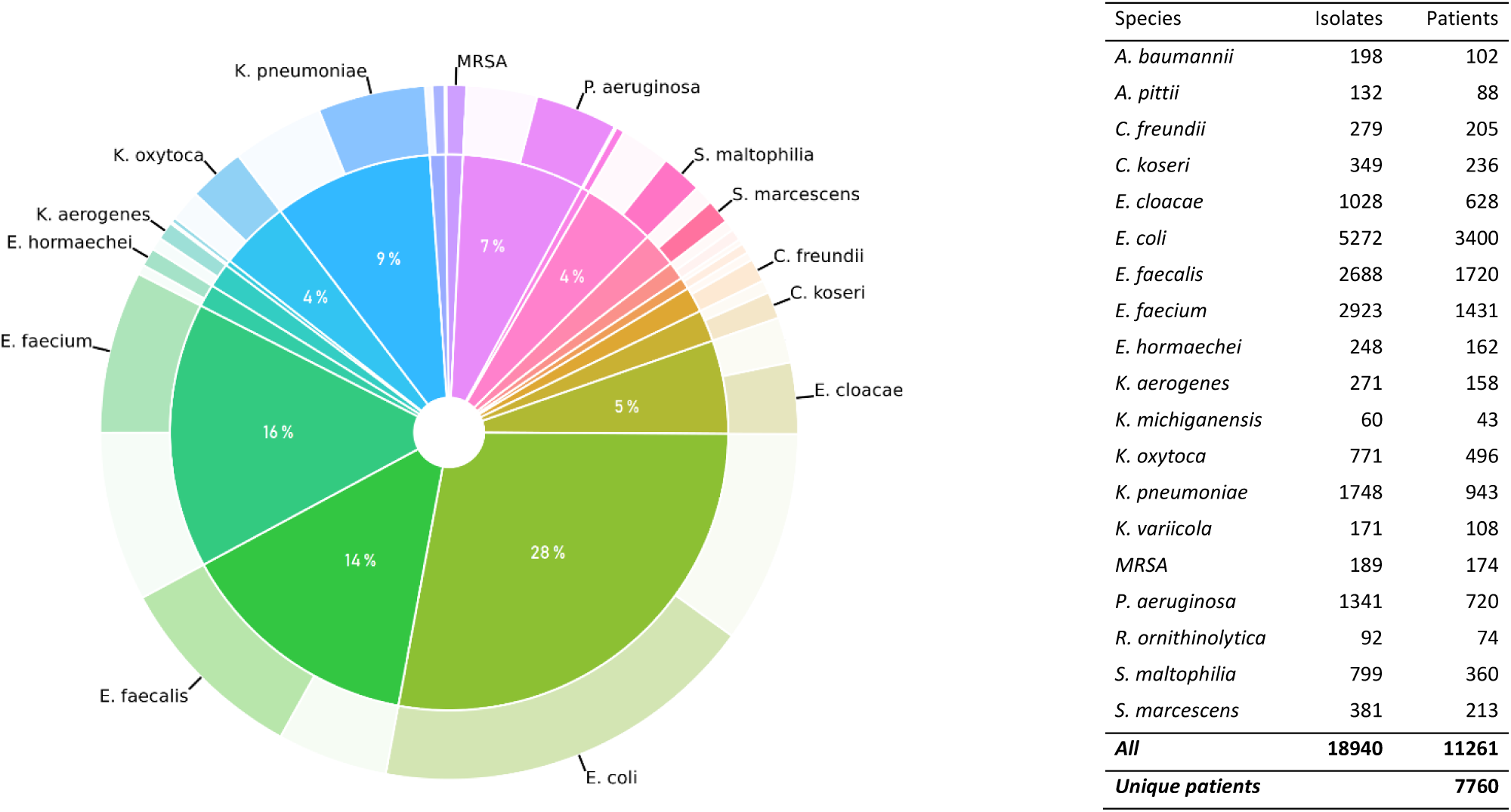
Relative distribution of bacterial species among the clinical isolates and corresponding patients from whom they were isolated. The figure illustrates the relative distribution of clinical isolates by species and the distribution of patients carrying at least one of these isolates over the 28-month study period. **Inner circle:** shows the distribution of clinical isolates (18 940) included in this study according to their species affiliation. **Outer circle:** represents fraction of individual patients from whom at least one isolate of the respective species was isolated (7760). The bacterial species included in this study are listed on the right, the absolute numbers of sequenced bacterial of the respective species are depicted in addition to the number of patients from whom the isolates were recovered.

### Identification of epidemiological links between patients harbouring genetically related clinical isolates

We found that overall, 27·1 % of all patients with at least one bacterial culture processed and analysed by WGS at the DCM harboured an isolate genetically related to an isolate from another patient. 583 isolates (from overall 551 patients) were MDROs with a genetic distance of 20 SNPs or less from a bacterial isolate found from another patient (Figure 2). In comparison, 1738 isolates (from overall 1552 patients) were non-MDROs with a genetic distance of 20 SNPs or less from a bacterial isolate found from another patient. Among the patients with a genetically linked MDRO, 385 and 76 had direct and indirect patient contact, respectively. For the patients with a genetically linked non-MDRO, 748 patients had direct and 247 indirect patient contact. In total, a direct or indirect epidemiological link was identified in 69 % of all patients, who harboured a genetically related clinical isolate.

**Figure 2:**
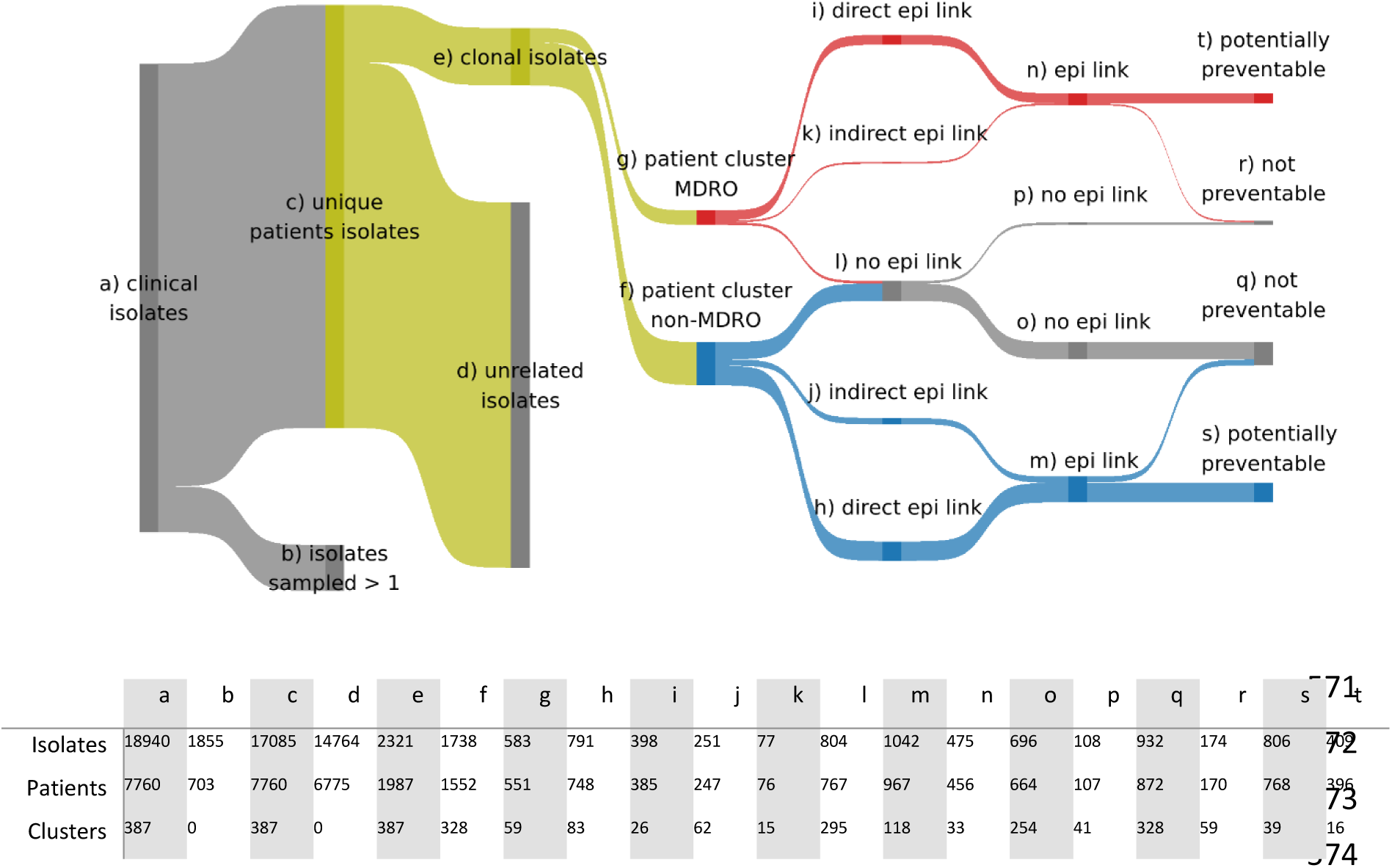
Sankey diagram depicting the distribution of patients, who harbor genetically related MDROs and non-MDROs, with and without the identification of direct and indirect epidemiological links. **a)** *Clinical isolates:* 18 940 clinical isolates were subjected to WGS within the 28-month study period. **b)** *Isolates sampled >1:* 1 855 of those were clonal variants that were repeatedly isolated from the same patient and thus excluded from further analysis. **c)** *Unique patient isolates*: 17 085 clinical isolates were unique patient isolates that were isolated from overall 7 760 patients. **d)** *Unrelated isolates*: A total of 14 764 isolates were genetically distinct, differing by more than 20 SNPs. **e)** *Clonal isolates*: 2321 isolates were genetically related to at least one other isolate (less than 20 SNP difference). **f)** *Patient cluster, non-MDRO:* 1552 patients were found in clusters harboring the same closely related non-MDRO. **g)** *Patient cluster, MDRO*: 551 patients were found in clusters harboring the same closely related MDRO. **h)** and **i)** *Direct epi link*: in 748 and 385 patients harboring non-MDRO and MRDO isolates, respectively, a direct epidemiological link was identified. **j)** and **k)** *Indirect epi link*: in 247 and 76 patients harboring non-MDRO and MRDO isolates, respectively, an indirect epidemiological link was identified. **l)** *No epi link*: 804 isolates had no identified epidemiological link in the hospital. **m)** *Epi link*: 967 non-MDRO and **n)** 456 MDRO harboring patients with either a direct or indirect epidemiological link, **o**) No epi link: 696 isolates from non-MDRO clusters and **p)** 108 isolates from MDRO clusters belonged to patients with no identified epidemiological link. **q)** 932 isolates from non-MDRO patients and **r)** 174 isolates from MDRO patients would not have been preventable because no link could be found or the corresponding patients were the 1^st^ or 2^nd^ cases within the clonal cluster. Assuming that targeted IPC measures have the potential to prevent further transmission after the identification of a genetically related isolate in a second patient, this means that transmission **s)** 806 non-MDRO transmissions and **t)** 409 MDRO transmissions could potentially be prevented because the respective patients had a traceable epidemiological link and were at least the 3^rd^ identified and epi linked candidate in the respective clonal cluster.

The likelihood of acquiring a bacterial isolate with an epidemiological link to another patient’s isolate was significantly higher for certain bacterial species. Most hospital transmissions, regardless of the resistance profile, involved *E. faecium* (Figure 3). Epidemiological links were also frequently identified for MRSA and *P. aeruginosa*. Of note, albeit the overall number of *E. coli* isolates with similar genetic profiles found in different patients was high, the epidemiology showed that most *E. coli* acquisitions could not be traced back to a transmission within the hospital. This was true for both resistant and, especially, for susceptible isolates, the latter of which were far more frequent (Figure 3). Additionally, many transmissions involved *P. aeruginosa* and *E. faecalis* isolates, although no transmissions of MDROs of these two species were detected. This suggests that previous studies focusing solely on MDRO transmission underestimated the spread of these two bacterial species in the hospital.

**Figure 3:**
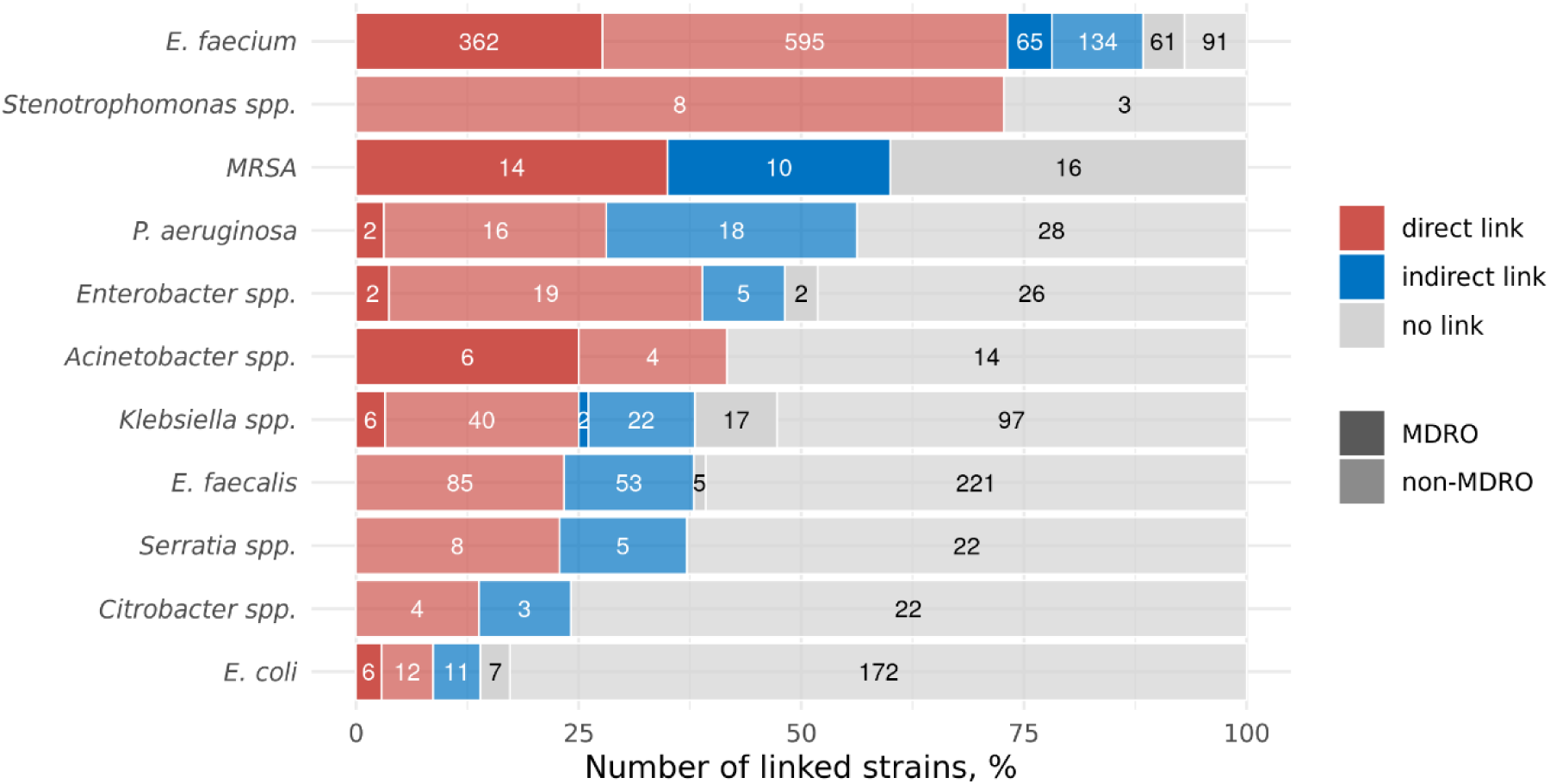
Epidemiological links between patients harbouring genetically related bacterial isolates. The relative fraction of direct (red), indirect (blue) and no epidemiological links (grey) of the patients harbouring phylogenetically closely related isolates of the indicated bacterial species for MDRO clusters (dark coloured) and non-MDRO clusters (light coloured) are depicted.

### Number and sizes of patient clusters harbouring genetically related clinical isolates

The 583 genetically linked MDRO isolates were obtained from overall 551 patients, who grouped into 59 clusters. Of these 551 patients, 456 MDRO-positive patients had an identified epidemiological link (Figure 2 and 4A). While the majority of cluster were small, the majority of patients were found in very large clusters. In particular, there were large differences in the distribution of cluster sizes among the bacterial species, with *E. faecium* clearly standing out and being consistently associated with large clusters. MRSA and MDR *Klebsiella spp*. were clearly in second and third place.

**Figure 4:**
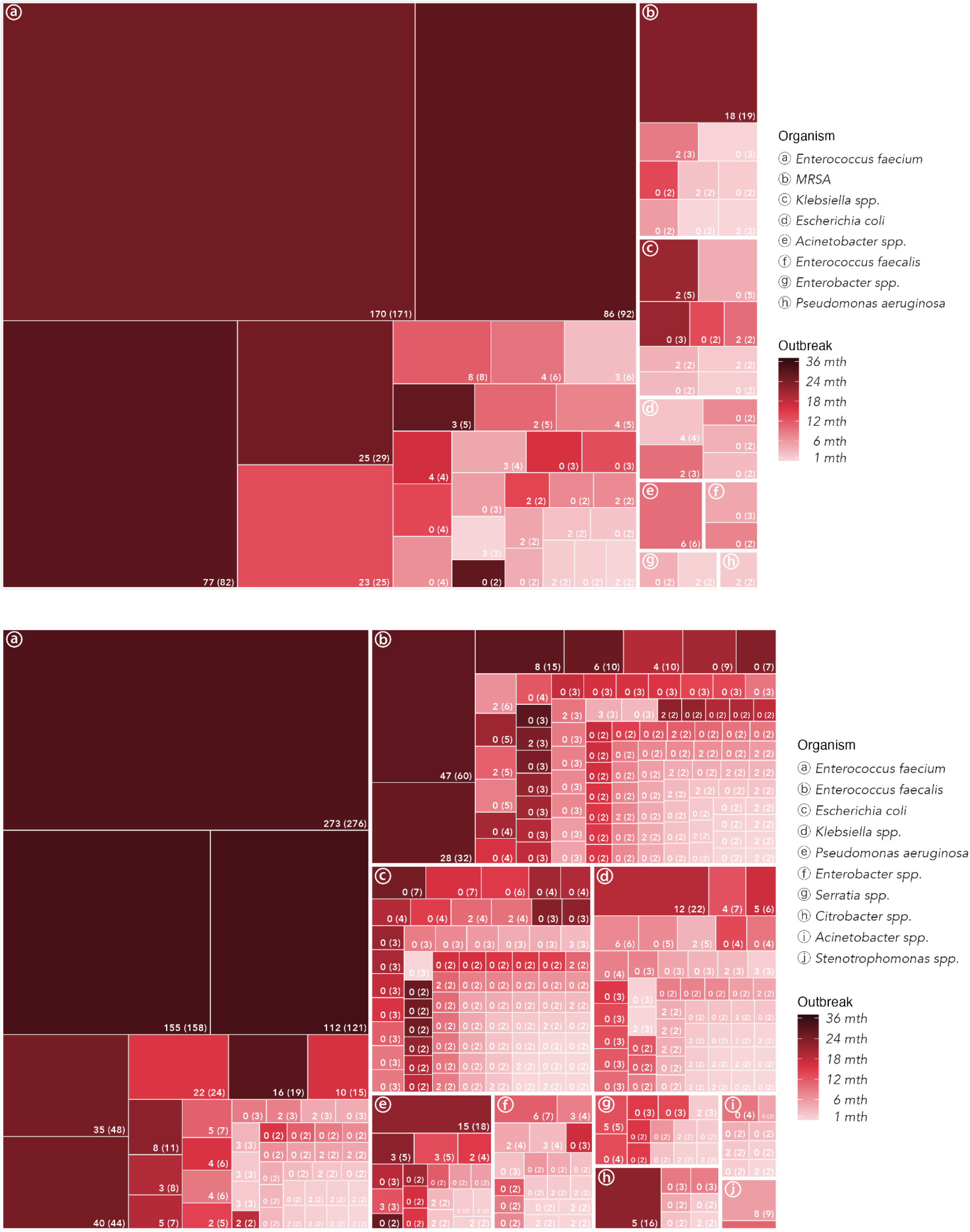
Number and sizes of patient clusters harbouring genetically related clinical isolates. **(A)** A total of 551 patients grouped into 59 clusters of patients from whom a closely related MDRO isolate was recovered. **(B)** A total of 1552 patients grouped into 328 clusters of patients from which the same genetically related non-MDRO isolate was recovered. The number of direct or indirect epidemiological links between the respective patients is depicted as well as the overall number of patients within the cluster (in brackets). The color code indicates the time between the recovery of the first and the last isolate of the respective cluster. The clusters were defined as a MDRO clonal cluster, if at least 90% of the clinical isolates of the respective patient cluster were categorized as MDROs.

In comparison, the 1738 non-MDROs isolates with a genetic distance of less than 20 SNPs from another patient’s isolate came from 1552 patients, who grouped into 328 clusters. For 967 of these patients a direct or indirect epidemiological link was identified (Figure 2 and 4B). Not only did *E. faecium* stand out, but also *E. faecalis*, both of which were associated with very large and large clusters, respectively. The cluster sizes of the other bacterial species were much smaller, with *E. coli, Klebsiella spp.* and *P. aeruginosa* topping the list. In general, and as expected, the time between the recovery of the first and last isolate of the respective clusters was longer for larger clusters and often stretched over years.

### Identification of local hotspots of transmission

For the six example pathogens—*K. pneumoniae, P. aeruginosa, E. coli, E. faecalis, E. faecium,* and MRSA—we further analyzed whether there is a preferred site of transmission within the hospital and if this varies by pathogen. As shown in Figure 5, the transmission rates of different pathogens varied significantly across wards. *K. pneumoniae* transmission was frequently observed in the neonatal units during our 28-month observational study. Similarly, *E. coli* and MRSA were also commonly transmitted among neonates. Furthermore, *E. faecium* exhibited a notable proportion of transmissions in the hematology department, while nearly half of the *P. aeruginosa* transmissions occurred among patients in intensive care.

**Figure 5:**
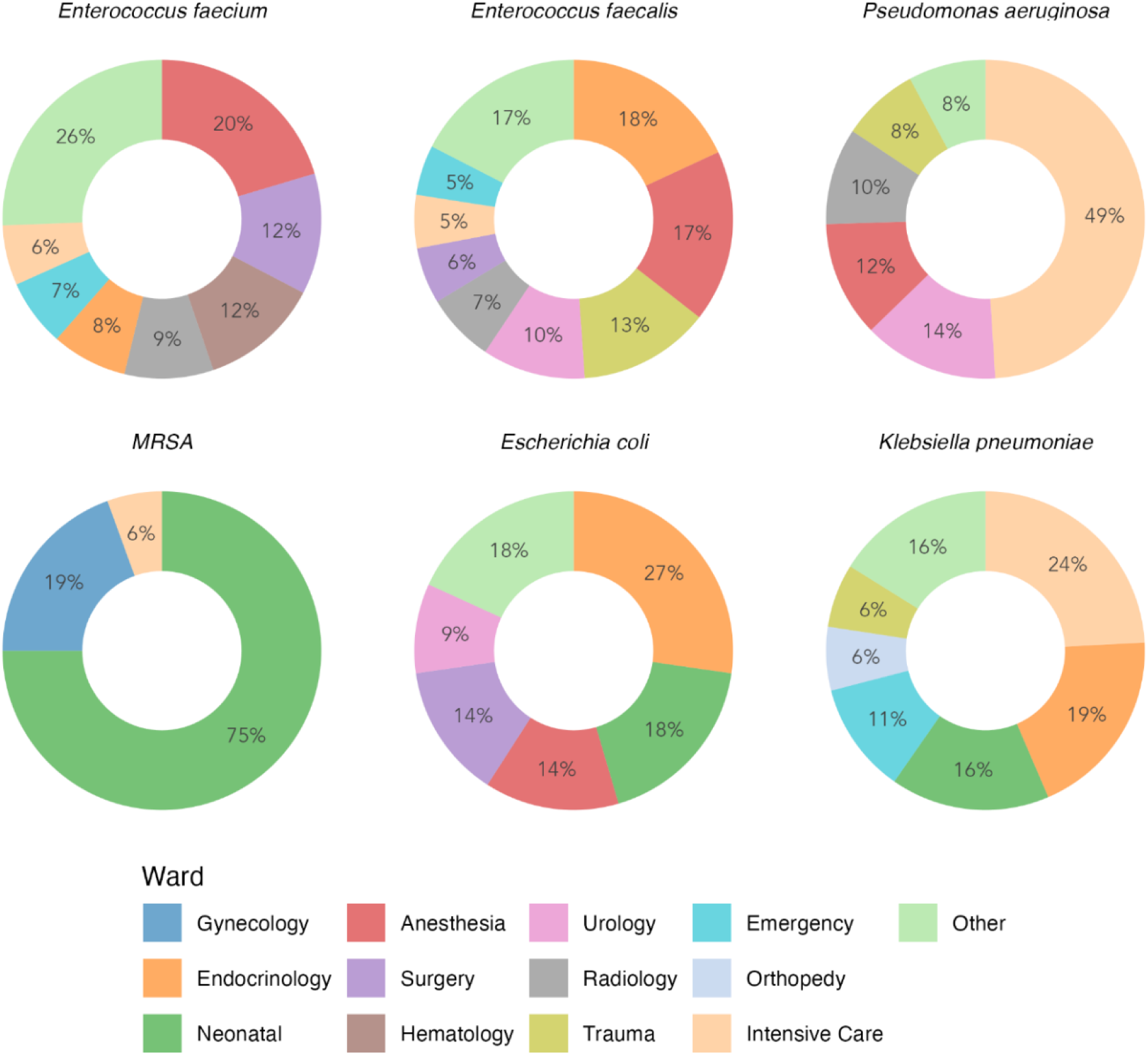
Identification of local hotspots of transmission for six dominant opportunistic pathogens during the 28-month observational study.

### Potential Impact of Genomic Epidemiology on cost-effectiveness

In our cost-effectiveness and clinical utility analysis, we assumed that the chain of transmission can be broken when WGS data is acted upon immediately. Effective environmental cleaning, patient isolation, and contact tracing were presumed to prevent pathogen transmission in all cases. The number of patients in whom infections or colonization could have been prevented was calculated after WGS identified a cluster of patients and infection control measures were initiated to interrupt the transmission route following the second transmission occurrence. Based on these assumptions, our data from the 28-month observational period suggest a potential reduction of 409 MDRO transmissions. Considering that 27·4 % of the MRDOs were identified in screens and not in clinical samples, we estimated the reductions in costs associated with MDRO colonization was €71 800 annually and reduction in costs associated with infection was €645 200 annually. If WGS was implemented solely for MDRO surveillance, the estimated annual costs of WGS would be €71 800. This would result in net cost savings of €645 600 annually for the implementation of genomic epidemiology for MDRO at Rigshospitalet.

Genomic epidemiology does not only benefit MDRO cases but also cases with transmission of susceptible, non-MDRO bacteria. Assuming genomic epidemiology enables to interrupt the transmission route after the second occurrence of the transmission of a susceptible isolate (non-MDRO), a total of 806 transmissions could have been prevented. This prevention amounts to an absolute cost reduction of approximately €875 700 annually, if proactive WGS was implemented at Rigshospitalet for the most important susceptible HAI-associated pathogens. However, extending WGS to also include susceptible isolates would cost, an additional €212 700 annually. Figure 6 illustrates the changes in costs and savings per year.

**Figure 6:**
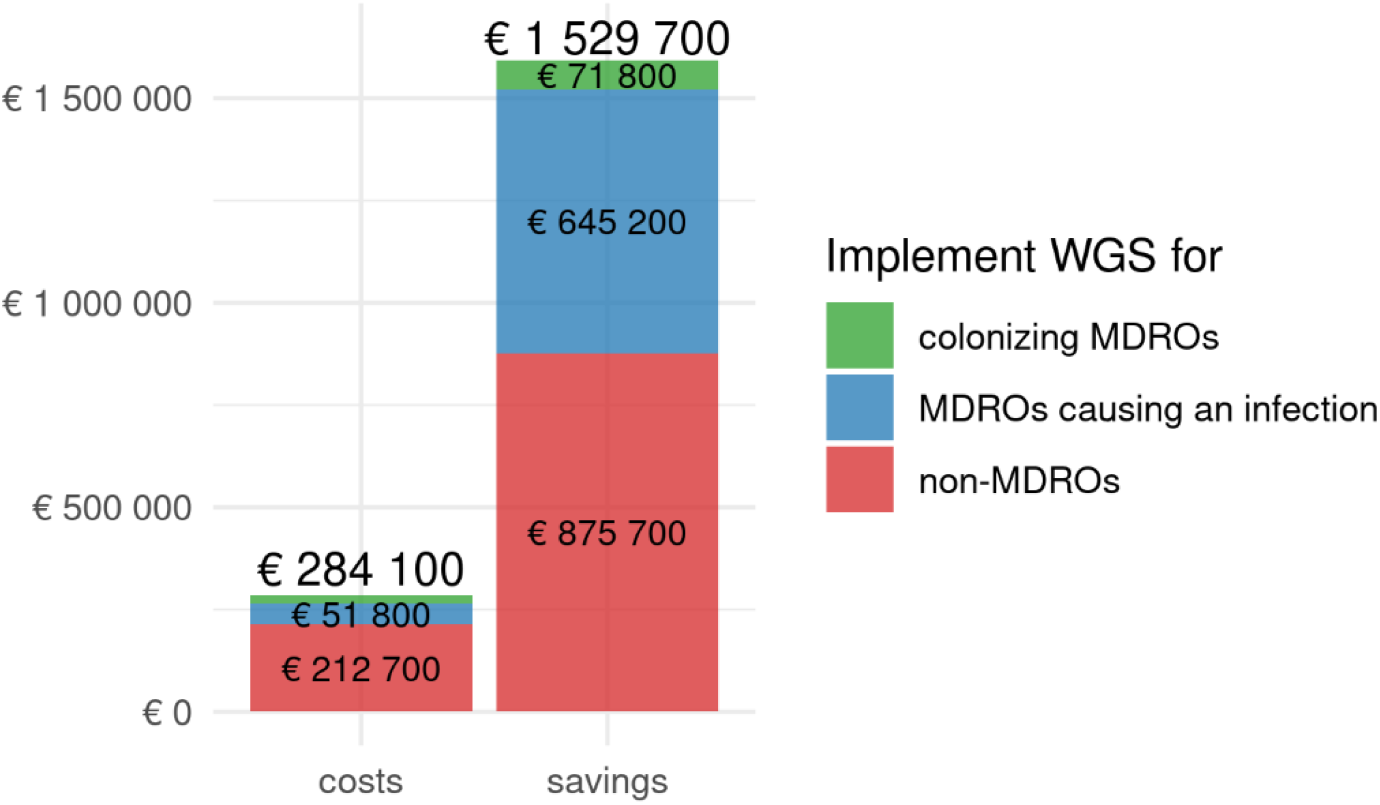
Overall annual potential savings and costs by use of the proactive genomic epidemiology approach. The health economic assessment uncovered yearly net cost savings of approximately € 1·25 million if initiated IPC measures completely halt transmission of clinical isolates after the second patient. The reduction in costs is due to a lower rate of costs for non-MDRO (red) and MDRO (blue) infections, respectively as well as colonization with MDRO (green). The costs for non-MDRO infections (red) are made up of the additional days a patient has to spend in hospital due to the infection (7·8 days), multiplied by the costs per patient for an additional day in hospital (€325). The costs for the colonization with MDRO (green) is composed of the average number of days a patient spends in the hospital (4·6 days), multiplied by the extra-costs incurred per patient per day for the isolation of the patient (€325). The cost for MDRO infections (blue) results from the average number of days a patient with an MDRO infection has to spend in hospital (7·8 days) multiplied by the additional costs incurred per day by a patient with an MDRO infection (€650).

### Potential Impact of Genomic Epidemiology on clinical utility

Instead of examining only the financial costs associated with implementing WGS, the burden of disease in a population should also be considered. Estimates for the burden of HAIs vary widely between sources, indicating a lack of consensus on the disease’s impact on quality of life. Since HAIs are typically short-term conditions, calculating Quality-Adjusted Life Years (QALY) for such a brief period can result in only a relatively small adjustment. A recent study addressed the challenge of calculating the burden of common HAIs (6). According to their calculations, a case of health care associated (HA) pneumonia resulted in 2·2 DALYs, HA blood stream infections (BSI) in 8 DALYs, HA urinary tract infections (UTI) in 0·8 DALYs, surgical site infection (SSI) in 0·5 DALYs and HA neonatal sepsis in 12·1 DALYs per case. We analysed the incidence of the different most common HAIs and assigned DALYs to the 297 MDRO (excluding the 112 MDRO colonisation cases) and 806 non-MDRO infections identified as preventable during the 28-month study period. In this way, we found that the introduction of genomic epidemiology could lead to savings of 1 818 DALYs over the 28-month study period (779 DALYs annually), with 75% of the DALYs from the most common HAIs (6) attributed to premature death (YLL) and 25% to years lost due to disability (YLDs)."

### Incremental cost-effectiveness ratios (ICERs) for guiding WGS resource allocation

We then evaluated whether the up-front investment in WGS for bacterial pathogens varies in cost-benefit depending on the bacterial species being sequenced, and identified which species offer the greatest financial advantage for the hospital. For several species, the investment in obtaining WGS did not yield any returns. For species, such as *E. coli, P. aeruginosa,* and *K. pneumoniae*, the initial investment did not break even. In contrast, for species like *E. faecium*, MRSA, *A. baumannii* and *E. faecalis* the investment resulted in net savings. For *E. faecium*, the savings were as high as €32 for every €1 invested (Figure 7). This is certainly due to the finding that *E. faecium* occurs in large outbreaks and therefore significantly greater savings can be achieved here by preventing the outbreak at an early stage. Even though in this study, for example, more was paid for the sequencing of *E. coli* than could be saved in costs by preventing an *E. coli* outbreak, this does not mean that sequencing *E. coli* might not be financially worthwhile in the long term. Our data show that *E. coli* is likely transmitted, but not necessarily in hospitals, as we found relatively few epidemiological links, particularly in MDRs, despite the genetic relatedness identified. The introduction of genomic surveillance that includes *E. coli* isolates outside the hospital could provide valuable new insights into transmission routes in the future.

**Figure 7:**
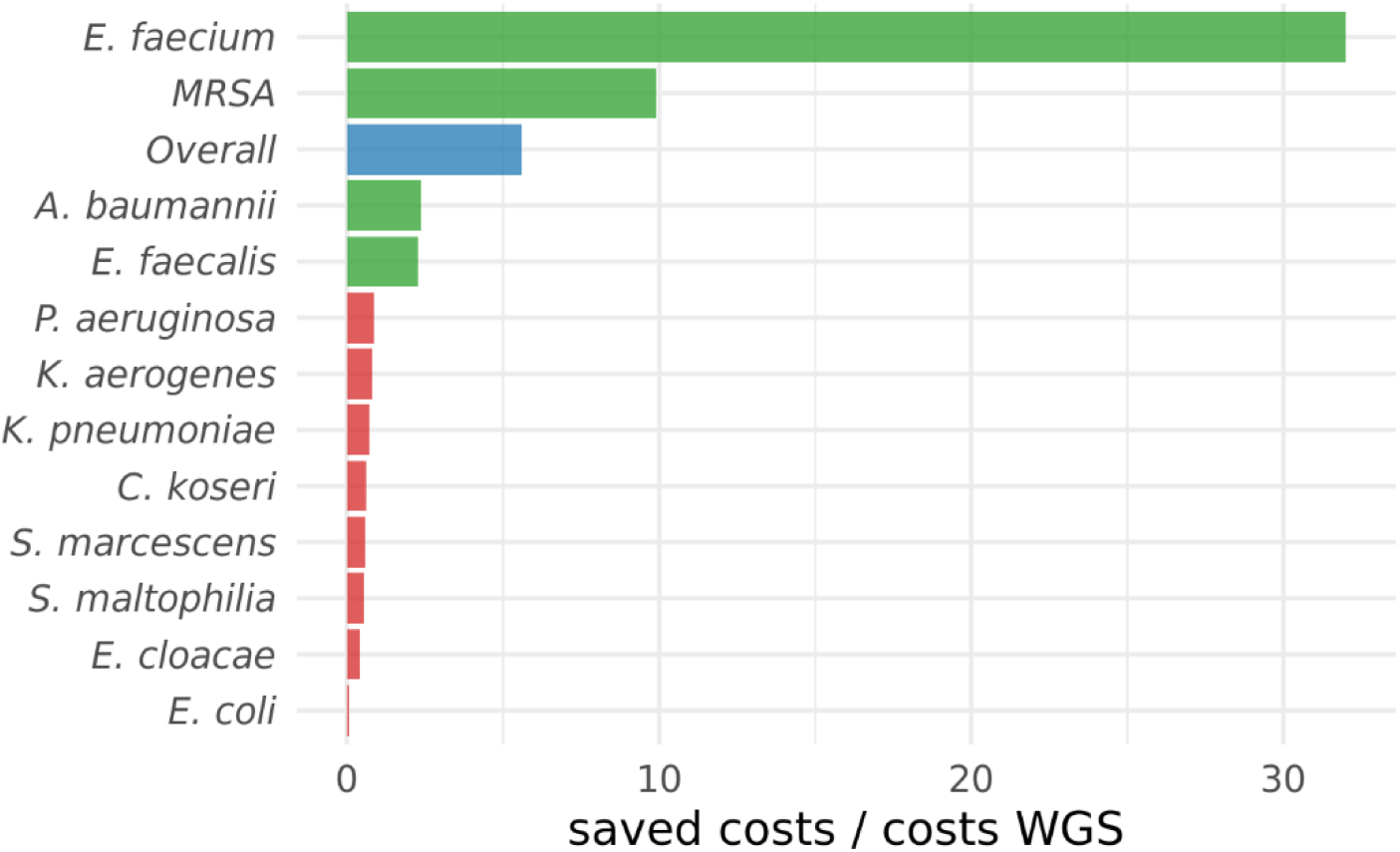
Net savings per investment in WGS of different bacterial species. Cost of WGS / saved costs is depicted for the implementation of a proactive WGS approach of all bacterial species included in this study (overall) and individually for each of the most frequent bacterial species, listed on the left. Bacterial species for which the initial investment in WGS does not break even (cost of WGS / avoided cost is < 1) are highlighted in red.

We also calculated the number of clinical isolates overall and within each species that need to be sequenced before a genetically linked second isolate can be identified (Figure 8). Due to the large clusters of *E. faecium* infected patients, it is not surprising that the number of clinical *E. faecium* isolates required to find a genetically linked isolate in another patient was low (under five). However, the numbers of MRSA, *E. faecalis* and *A. baumannii* isolates that needed to be sequenced before a clonally related isolate was detected were also under 25. Thus, although fewer isolates were identified for these species overall in absolute numbers, their relative contribution to outbreaks was high.

**Figure 8:**
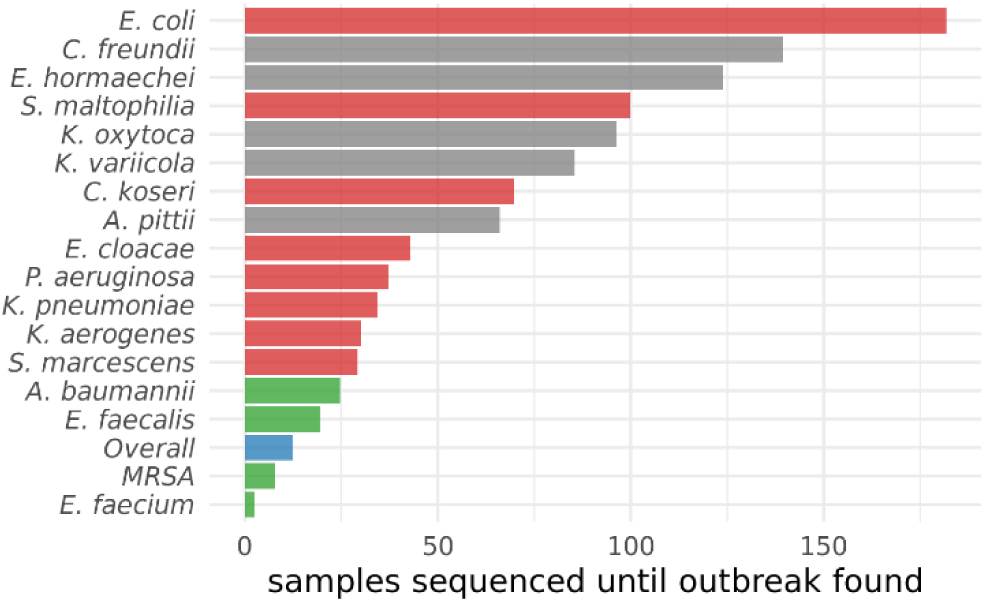
Number of genomes needed to be sequenced before a genetically linked second isolate is identified. The number of clinical isolates that needed to be sequenced before a genetically linked second isolate was identified is depicted for the implementation of a proactive WGS approach of all bacterial species included in this study (overall) and individually for each of the most frequent bacterial species, listed on the left. Bacterial species for which the initial investment in WGS does not break even (cost of WGS / avoided cost is < 1) are highlighted in red (coloring same as in Figure 6).

**Figure 9:**
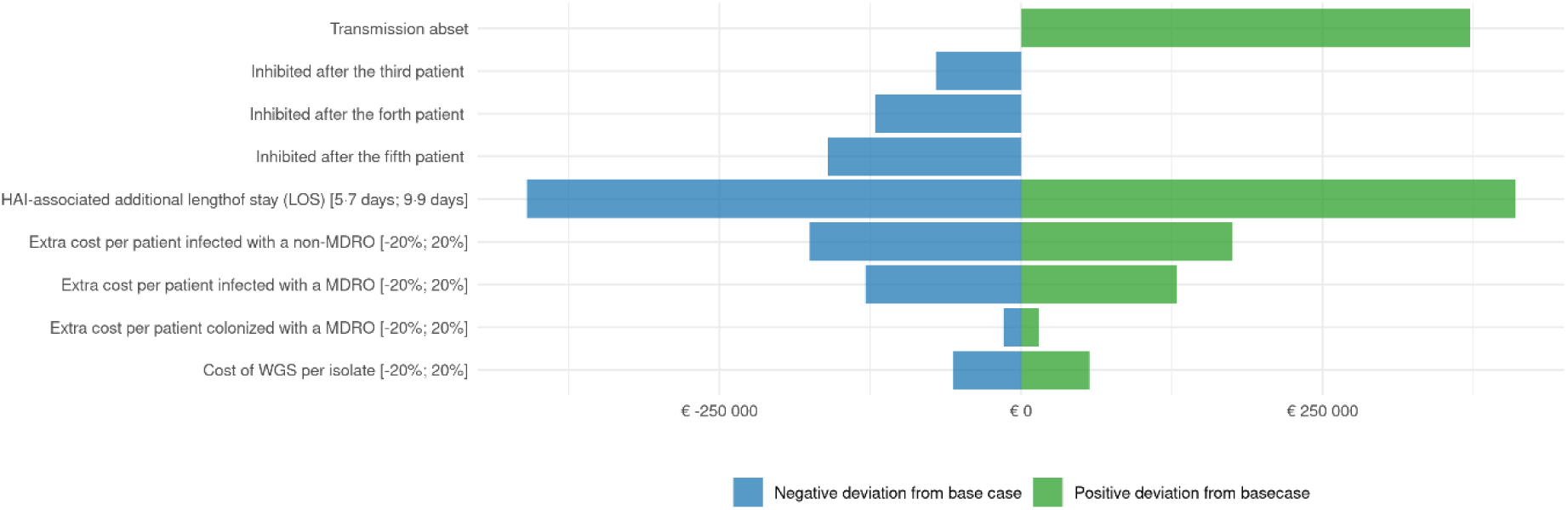
Sensitivity analysis on hospital savings. Comparing costs of hospital care: plausible alternative values to the base case in terms of how many transmissions are detected before the outbreak is stopped, additional length of stay, extra costs of hospital stay or MDRO colonization and cost of WGS as listed in Table 1 (95% confidence interval or +/- 20% of the base case costs) were used in the sensitivity analysis and differences in net hospital cost savings are depicted as deviations from the base case.

### Sensitivity analysis

A sensitivity analysis was used to illustrate and assess the confidence level of our economic evaluatiońs conclusions. The analysis showed that even when plausible alternative values (95% confidence interval or +/- 20% of the base case costs) were applied, hospital cost savings were always consistently retained. The results were most sensitive to changes in the extra length of hospital stay due to the acquisition of an infection. However, even if the base case of an additional hospital stay was reduced from 7·8 days to 5·7 (lower bound of the 95% CI of 5·7-9·9) (13), the potential cost savings still amounted to overall € 899 097 annual saving for the hospital. The sensitivity analyses also demonstrate that if there were no transmissions at all, the hospital would save overall € 1 680 657. If transmission was only stopped after the third patient (instead of after the second patient as in the base case), cost savings would be reduced to € 1 238 139. Stopping transmission after the fourth patient would lead to savings of € 1 187 769 and after the fifth patient of € 1 148 757. This indicates that a significant portion of the cost-benefits come from curtailing large outbreaks.

The low base case costs for WGS of €35/sample at Rigshospitalet strongly depend on the number of samples processed in parallel, as more WGS results in a lower cost per sample. Raising the WGS costs shows a breakeven point at €196/sample, after which higher WGS costs will result in a net cost for the hospital. Figure 8 illustrates the different costs used in the estimations as well as the variations in net cost results compared to the base case when implementing these changes.

## Discussion

Increasing evidence suggests that genomic epidemiology is superior for confirming or refuting transmissions and outbreaks of numerous hospital pathogens (15). When performed routinely rather than retrospectively, it can inform standard IPC investigations. This helps detect and contain outbreaks that traditional IPC methods might miss, identify transmission routes, and correct misidentified outbreaks where no transmission occurred. The potential benefits include enhanced patient safety and substantial cost savings through a more targeted use of IPC time and resources, focusing on true outbreaks and de-escalating clusters that occurred by chance (16). However, despite many initial studies suggesting these advantages (16), clinical evidence for improvements in clinical outcomes and hospital budgets are still lacking (29). Studies have been limited to a specific selection of pathogens and hospitals and were based on extrapolated assumptions from partial WGS data.

In this study, we expanded the focus of genomic epidemiology to a broad spectrum of pathogens. We performed WGS on 19 gram positive and gram negative pathogen species, covering approximately 33·5 % of all bacterial isolates recovered from one patient per day and isolation site at the Department of Clinical Microbiology of a large university hospital in Denmark over 28 months. We also extended the WGS strategy to include non-MDROs. By expanding genomic epidemiology to a wider range of pathogens, and by including non-MDRO, we aim to build a comprehensive understanding of pathogen dynamics in the hospital environment and to inform better IPC practices and policies that can adapt to various pathogen threats.

Our holistic approach shed light on the previously unexplored transmission landscape across the entire hospital. By subjecting isolates to WGS irrespective of their site of isolation, the ward in which the patient was hospitalized, and their antibiotic susceptibility profile, we found that 27·1 % of all hospitalized patients with at least one positive bacterial culture, harboured a bacterial isolate also found in another patient. For 69 % of the patients harbouring a closely related bacterial isolate an epidemiological link was identified in the hospital. The great majority of possibly transmitted isolates were non-MDROs, with 2·2 times more transmissions of antibiotic-susceptible isolates than resistant ones. This suggests that earlier studies on MDRO transmission were only the tip of the iceberg and highlights a significant problem in hospitals: transmission is widespread and not limited to cases involving resistant isolates. Interestingly, despite the lower potential cost savings from preventing infections caused by non-MDROs and the significantly higher investment in WGS for susceptible isolates, monitoring non-MDROs resulted in greater overall cost savings for the hospital compared to monitoring only MDROs.

Notably, Enterococci, including antibiotic susceptible isolates, were transmitted particularly frequently, suggesting that they can more easily overcome current hospital hygiene practices than other species (30). Future efforts should focus on investigating the circumstances of enterococcal transmission in hospitals, identifying environmental and patient-related factors, and examining clinical care practices in the management of patients that may contribute to the spread of these pathogens. By understanding these factors, we might be able to better target hygiene practices to contain the spread of enterococci and other resilient bacteria.

The reported transmission frequencies for individual pathogens should be interpreted with caution. The use of a general 20-SNP cut-off between isolates may influence the proportion of positives within each species, as species with high genetic diversity may require a higher SNP cut-off to detect meaningful patterns of relatedness. Conversely, species with lower genetic diversity are more likely to form larger clusters at the same 20 SNP cut-off. This could lead to an overestimation of transmission for some bacterial species and an underestimation for others. Additionally, the frequency of sampling impacts transmission detection—greater WGS sampling (including screens) increases the likelihood of identifying genetically related isolates and epidemiological links. As a result, the transmission rate reported in this study—27·1% of patients carrying a clinical isolate genetically related to related to another patient’s—may increase with more intensive sampling. WGS surveillance could reveal significantly higher transmission rates, particularly for those species that were sampled less frequently. This is concerning, highlighting the need for future studies to refine transmission estimates and evaluate whether WGS-informed IPC measures can effectively reduce transmission rates in settings where transmission is frequent.

Interestingly, we found many *E. coli* isolates with close genetic relatedness but no epidemiological link in the hospital. This has been observed before (31) and was true for resistant but especially for susceptible isolates. Detecting non-hospital associated sources of clonally related *E. coli* isolates will be crucial in the future. To achieve this, it will be necessary to combine WGS strategies in hospitals with those in outpatient facilities and residency homes to track transmission at a higher resolution to detect missing links. This approach will help to identify transmission routes, and host-, bacteria-, environment- and patient care-associated factors that facilitate spread of *E. coli* and possibly other Enterobacterales. This promises to safeguard patients as those pathogens are increasingly contributing to the infectious disease burden, particularly in vulnerable patients.

We furthermore demonstrate that despite the substantial up-front investment required for routine WGS, implementing a genomic epidemiology approach in the hospital holds significant potential for cost savings that remain largely insensitive to variations in input parameters. However, we observed considerable variations in the sizes of infection clusters depending on the pathogen. This impacts the effectiveness of a general WGS-based surveillance strategy, as substantial cost savings were primarily seen with a limited range of pathogens, mainly enterococci. For other selected pathogens, the upfront investment did not pay off. In an era of tight hospital budgets, these findings could help guide the allocation of WGS resources. Nevertheless, our results also show that sequencing a broader range of pathogens at least break even in terms of costs. It is important to recognize that, in the long run, a general proactive WGS-based approach may be beneficial and cost-effective. Expanding proactive WGS to identify additional transmission pathways and missing links beyond the hospital could further inform and tailor infection control measures to real-life situations (32). Finally, it is noteworthy that implementing a WGS-based surveillance potentially could save more than 750 DALYs per year at Rigshospitalet; a significant reduction that should be highly valued.

In conclusion, our findings clearly demonstrate that broad genomic epidemiology has significant potential to improve patient care while being cost-effective. Future studies should evaluate whether proactive implementation of genomic epidemiology in hospitals consistently leads to a reduction in the incidence of HAIs and substantial cost savings across diverse healthcare settings. Even if the actual cost savings are lower than anticipated, widespread adoption of WGS could uncover previously unexplored transmission routes, refine general infection control strategies, and enable more targeted interventions. Expanding surveillance to residential homes and healthcare facilities outside the hospital could further reveal pathogen transmission dynamics, significantly contributing to better understanding of pathogen spread. This would be crucial for optimising IPC measures also in residential facilities and more broadly for public health.

## Data Availability

Data will be made available when submitted to a journal or by request.

## Author contribution

Conceptualization: Frederik Boëtius Hertz; Karen Leth Nielsen; Susanne Häussler.

Data curation: Frederik Boëtius Hertz; Karen Leth Nielsen; Martin Lucas Jørgensen, Theiss Bendixen, Roshkan Srinathan, Steen Christian Rasmussen; Rasmus L. Marvig; Christian Salgaard Jensen, Jelena Erdmann, Dmytro Strunin. Formal analysis: Martin Lucas Jørgensen, Theiss Bendixen, Roshkan Srinathan

Investigation: Frederik Boëtius Hertz; Karen Leth Nielsen; Martin Lucas Jørgensen, Theiss Bendixen, Roshkan Srinathan

Methodology: Frederik Boëtius Hertz; Karen Leth Nielsen; Martin Lucas Jørgensen, Theiss Bendixen, Roshkan Srinathan.

Ethical approval: Frederik Boëtius Hertz.

Resources: Frederik Boëtius Hertz; Karen Leth Nielsen; Susanne Häussler.

Software: Martin Lucas Jørgensen, Theiss Bendixen, Roshkan Srinathan.

Supervision: Susanne Häussler.

Project administration: Karen Leth Nielsen, Frederik Boëtius Hertz.

Writing – Original Draft Preparation: Frederik Boëtius Hertz and Susanne Häussler.

Writing – Review & Editing: Frederik Boëtius Hertz, Karen Leth Nielsen, Martin Lucas Jørgensen, Theiss Bendixen, Roshkan Srinathan, Steen Christian Rasmussen, Jelena Erdmann, Asger Nellemann Rasmussen, Dmytro Strunin, Christian Salgaard Jensen, Jenny Dahl Knudsen, Susanne Häussler.

Materials & Correspondence: Susanne Häussler.

Potential conflicts of interest: FBH, KLN, SCR, RLM, CSJ, JE, ANR, DS, JDK, SH declare no conflict of interest. MLJ, TB, RS were employed by Nordic Healthcare Group, Copenhagen, a private company during the study period.

Transparency declaration: All authors affirm that this manuscript is an honest, accurate, and transparent account of the study being reported and that no important aspects of the study have been omitted.

## Ethical approval

There were no ethical concerns for this study. This was an observational, non-intervention study. The extraction of data from registries was approved by the local data protection agency (Journal-nr.: R-21015888) and registered in Pactius (P-2020-743).

## Acknowledgement

We would like to thank Inken Häussler for her help in cost calculations and generating the figures.

